# Strain sharing and persistence of microbial pathogens colonizing the skin of residents in a regional nursing home network

**DOI:** 10.1101/2025.11.05.25339587

**Authors:** Yaovi M.G. Hounmanou, Gabrielle M. Gussin, Sean Conlan, Raveena D. Singh, Clay Deming, Diana M. Proctor, Marco Teixeira, Ashlee M. Earl, Colin J. Worby, Heidi H. Kong, Susan S. Huang, Julia A. Segre

**Affiliations:** Microbial Genomics Section, NHGRI, NIH, Bethesda, MD, USA; Division of Infectious Diseases, University of California Irvine School of Medicine, Irvine, CA, USA; Department of Microbiology and Molecular Genetics, The University of Texas Health Science Center at Houston, McGovern Medical School, Houston, TX, USA; Infectious Disease & Microbiome Program, Broad Institute, Cambridge, MA, USA; Delft Bioinformatics Lab, Department of Intelligent Systems, Delft University of Technology, Delft 2628XE, The Netherlands; Cutaneous Microbiome and Inflammation Section, NIAMS, NIH, Bethesda, MD, USA

**Keywords:** Skin Microbiome, Metagenome-Assembled Genomes, Multidrug Resistant Organisms, Antimicrobial Resistance in Nursing Homes

## Abstract

Antimicrobial resistance (AMR) is a global public health threat that disproportionately affects vulnerable populations, including nursing home (NH) residents. Surveillance and control in NHs are resource-limited and typically restricted to perirectal cultures, overlooking both skin colonization and multidrug-resistant organisms (MDROs) not recovered by selective media. Here we show, within the cluster-randomized Project PROTECT trial (NCT03118232), that residents’ skin serves as a major reservoir of transmissible MDROs. We analyzed 207 groin and axilla swabs from 38 residents across 15 California NHs using shotgun metagenomics, selective culturing, and isolate genome sequencing. Culture detected MDROs in 10 of 38 residents (26.3%), including extended-spectrum β-lactamase (ESBL)-producing *Escherichia coli* ST131/ST648 in 4 (10.5%) and methicillin-resistant *Staphylococcus aureus* in 7 (18.4%). In contrast, metagenome-assembled genomes identified broader MDRO colonization, including multidrug-resistant *E. coli* ST93 in 27 residents (71.1%), methicillin-resistant *Staphylococcus epidermidis* ST2 in 14 (36.8%), *Proteus mirabilis* in 16 (42.1%), *Providencia stuartii* in 7 (18.4%), *Enterococcus faecalis* in 7 (18.4%), and *Pseudomonas aeruginosa* in 5 (13.2%). Colonization persisted after bathing. Clonal *E. coli* ST93 (≤30 SNPs) was shared by 27 residents across 9 facilities, and 5 resident pairs (13.2%) carried clonally related strains of ≥2 MDRO species, consistent with polymicrobial transmission. Our findings demonstrated the skin as a persistent reservoir of MDROs and the importance of metagenomic surveillance to uncover hidden colonization and transmission pathways, underscoring the need to expand AMR monitoring in long-term care.

## Introduction

Antimicrobial resistance (AMR) threatens decades of medical progress, rendering once-treatable infections increasingly difficult or impossible to manage. Multidrug-resistant organisms (MDROs) colonizing and infecting patients undermine treatment efficacy, increase morbidity and mortality, and escalate healthcare costs. In 2021, an estimated 4.71 million deaths were associated with bacterial AMR, with global disability-adjusted life years projected to reach 46.5 million by 2050. ^1^ The economic toll of direct healthcare costs associated with AMR is estimated at 66 billion USD and predicted to reach 159 billion USD without improved interventions. ^2^ Older adults are disproportionately affected, with AMR-related deaths increasing by more than 80% in those aged 70 years and older between 1990 and 2021. ^1^ Many of these vulnerable individuals reside in nursing homes (NH). ^3^

Nursing homes are increasingly recognized as persistent reservoirs for MDROs. ^4^ In the USA, more than 1.3 million people live in 15,300 nursing homes, where MDRO prevalence is four to six times higher than in hospitals. ^3, 5^ Prolonged stays, frequent antibiotic use, hospital transfers, and close-contact communal living amplify opportunities for colonization and onward transmission. ^6^ Colonization is often asymptomatic but increases risk of infection, making detection essential for prevention. Most studies of MDRO carriage in NHs have relied primarily on culture-based detection from perirectal sites, providing only a partial view of MDRO ecology and transmission risk. ^7,8^ These approaches overlook skin colonization, fail to capture non-resistant or co-colonizing strains on selective media, and lack strain-level resolution of bacterial diversity and persistence.

Recent data show that the skin of nursing homes residents can harbor MDROs, including *Candida auris* and ESKAPE pathogens (*<u>E</u>nterococcus faecium, <u>S</u>taphylococcus aureus, <u>K</u>. pneumoniae, <u>A</u>. baumannii, <u>P</u>. aeruginosa, <u>E</u>nterobacter* species) ^9^. The inguinal crease and axilla are recognized hotspots for microbial colonization and potential transmission. ^7, 10, 11^ Yet, AMR surveillance focuses almost exclusively on perirectal sites and relies on culture-based methods that can miss co-colonizing strains, morphologically similar lineages, or organisms lacking expected resistance phenotypes. ^10^ Shotgun metagenomics analysis enables recovery of metagenome-assembled genome (MAGs) ^12^, providing strain-level resolution directly from complex samples. ^10,13^ Integrated genomic approaches combining culturomics and metagenomics now offer an unprecedented ability to track MDRO persistence and spread. Apart from our previous work^9^ applying shotgun metagenomics to NH skin colonization, no other study has used this approach, and none have combined metagenomics with bacterial isolate genome sequencing to study MDRO skin ecology. This study therefore advances the field by applying an integrated genomic approach to reveal the extent, persistence, and transmission dynamics of MDRO skin colonization in nursing home residents.

We leveraged the Project PROTECT trial, a cluster-randomized study of nursing homes in California, which demonstrated that MDRO colonization was highly prevalent in nursing homes and strongly associated with increased hospital transfers. ^5^ During this trial, skin swabs were collected from a convenience sample of residents for bacterial isolation and microbiome analysis. In this observational study, we integrated shotgun metagenomics and isolate sequencing to quantify MDRO colonization, persistence, and transmission among 38 residents in 15 nursing homes. We aimed to: (i) define the extent of MDRO skin colonization, (ii) assess strain persistence, (iii) characterize inter-resident and inter-facility strain sharing, and (iv) resolve the resistome structure. These findings illuminate cryptic, indolent skin colonization of nursing home residents with MDROs, informing potential future infection control strategies for long-term care settings.

## Results

### Selective culture reveals diverse MDROs on NH residents’ skin

Of the 38 residents across 15 NHs, selective culturing revealed 10 residents (26.3%) with skin MDRO carriage based on single timepoint sampling of the axilla and groin. Six residents (15.8%) carried ESBL organisms. Three residents (7.9%) in 3 different facilities carried *E. coli* ST131, a globally disseminated uropathogenic ESBL lineage, while ESBL-producing *E. coli* ST648, *Proteus mirabilis*, and *Providencia stuartii* were each recovered from a single resident. Vancomycin-resistant *Enterococcus faecalis* was recovered from 2 residents (5.3%), including ST109 and ST778 lineages (Table 1). In addition, methicillin-resistant *Staphylococcus aureus* (MRSA) was cultured from the skin samples of seven residents (18.4%). These findings confirm that skin can act as a reservoir for clinically relevant, antimicrobial-resistant organisms, with potential implications for infection risk and resident-to-resident dissemination.

**Table 1.** Skin-colonizing MDROs isolated from NH residents in this study.

| Species | MLST | #NHs | Residents Colonized | Phenotypic profile |
| --- | --- | --- | --- | --- |
| *Escherichia coli* | ST131 | 3 (20%) | 3 (7.9%) | ESBL |
|  | ST648 | 1 (6.7%) | 1 (2.6%) | ESBL |
| *Enterococcus faecalis* | ST109 | 1 (6.7%) | 1 (2.6%) | VRE |
|  | ST778 | 1 (6.7%) | 1 (2.6%) | VRE |
| *Proteus mirabilis* | NA | 1 (6.7%) | 1 (2.6%) | ESBL |
| *Providencia stuartii* | ST83 | 1 (6.7%) | 1 (2.6%) | ESBL |
| *Staphylococcus aureus* | - | 6 (40%) | 7 (18.4%) | MRSA* |
| Overall Skin MDRO carriage |  | 7 (46.7%) | 10 (26.3%) |  |

\*The MRSA isolates were not sequenced as part of in this observational study.

### Opportunistic pathogens persist within a dysbiotic skin microbiome

Analysis of the skin shotgun metagenomic data revealed a predominantly bacterial skin microbiome with typical commensal taxa (*Cutibacterium, Corynebacterium, Staphylococcus* spp.), surprisingly coexisting with opportunistic pathogens such as *Enterococci, E. coli, P. mirabilis, P. aeruginosa* and *Providencia* spp that are not expected in a normal skin microbiome. Fungal (*Malassezia* spp.) and viral taxa (Betapapillomavirus, Uroviricota phages) were consistently detected but at low relative abundances (Figure 1B).

**Figure 1.**
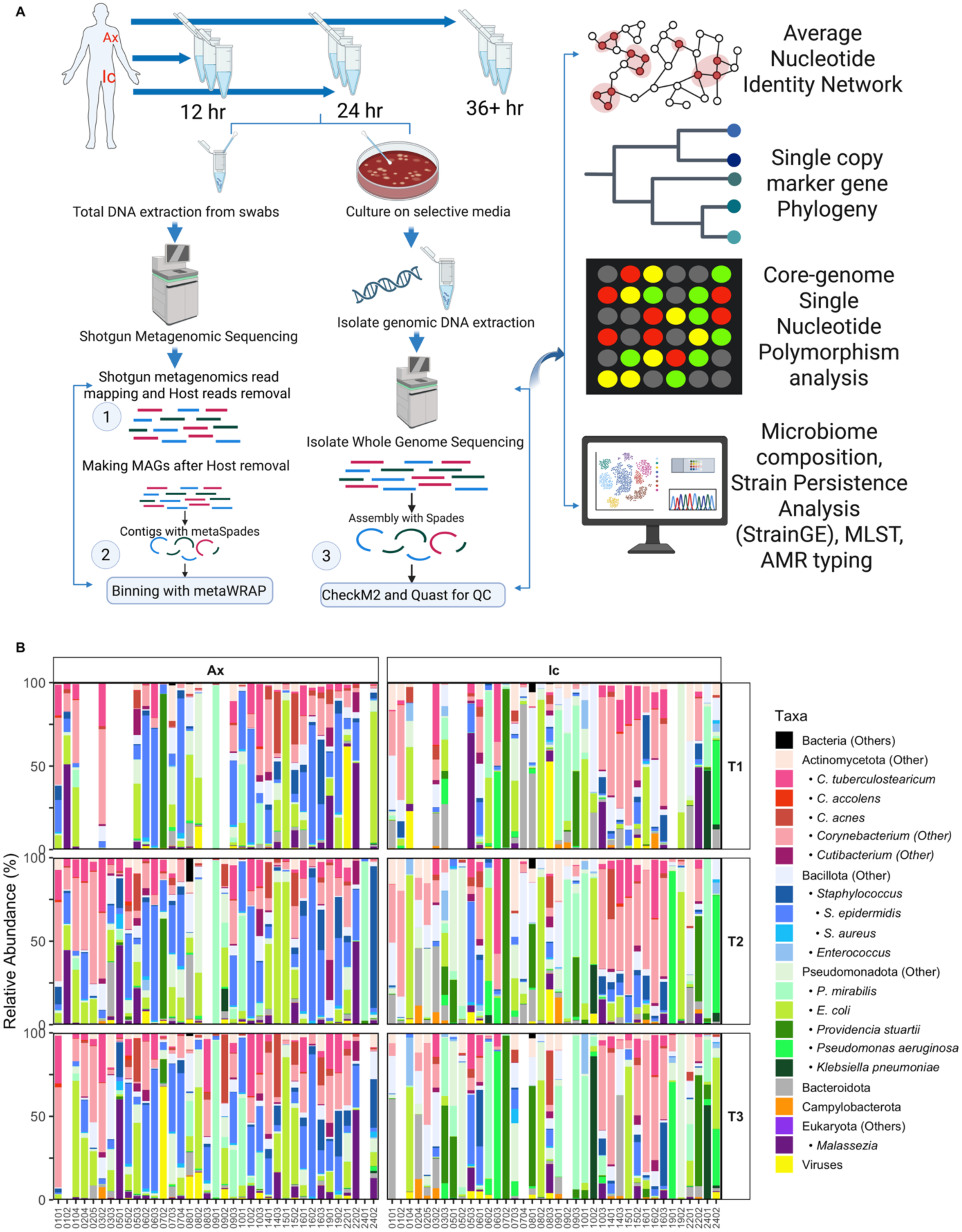
Overview of study design and skin microbiome composition analysis. (A) Study workflow for sample collection, sequencing, and genomic analysis of skin swabs collected from nursing home residents. (B) Stacked bar plots show the relative microbial abundances of individual nursing home residents, at two skin sites (axilla and inguinal crease) at three visits (T1, T2, T3).

Across samples, the overall composition of nursing-home residents’ skin microbiome appeared broadly similar between body sites and over time, with no significant differences detected in Shannon diversity between inguinal crease (Ic) and axilla (Ax) samples (p > 0.05). Descriptively, Enterobacterales reads (including *E. coli* and *P. mirabilis*) tended to be more prevalent in Ic samples, whereas axilla samples showed greater representation of Bacillota taxa such as *Staphylococcus epidermidis* (Figure 2B).

**Figure 2.**
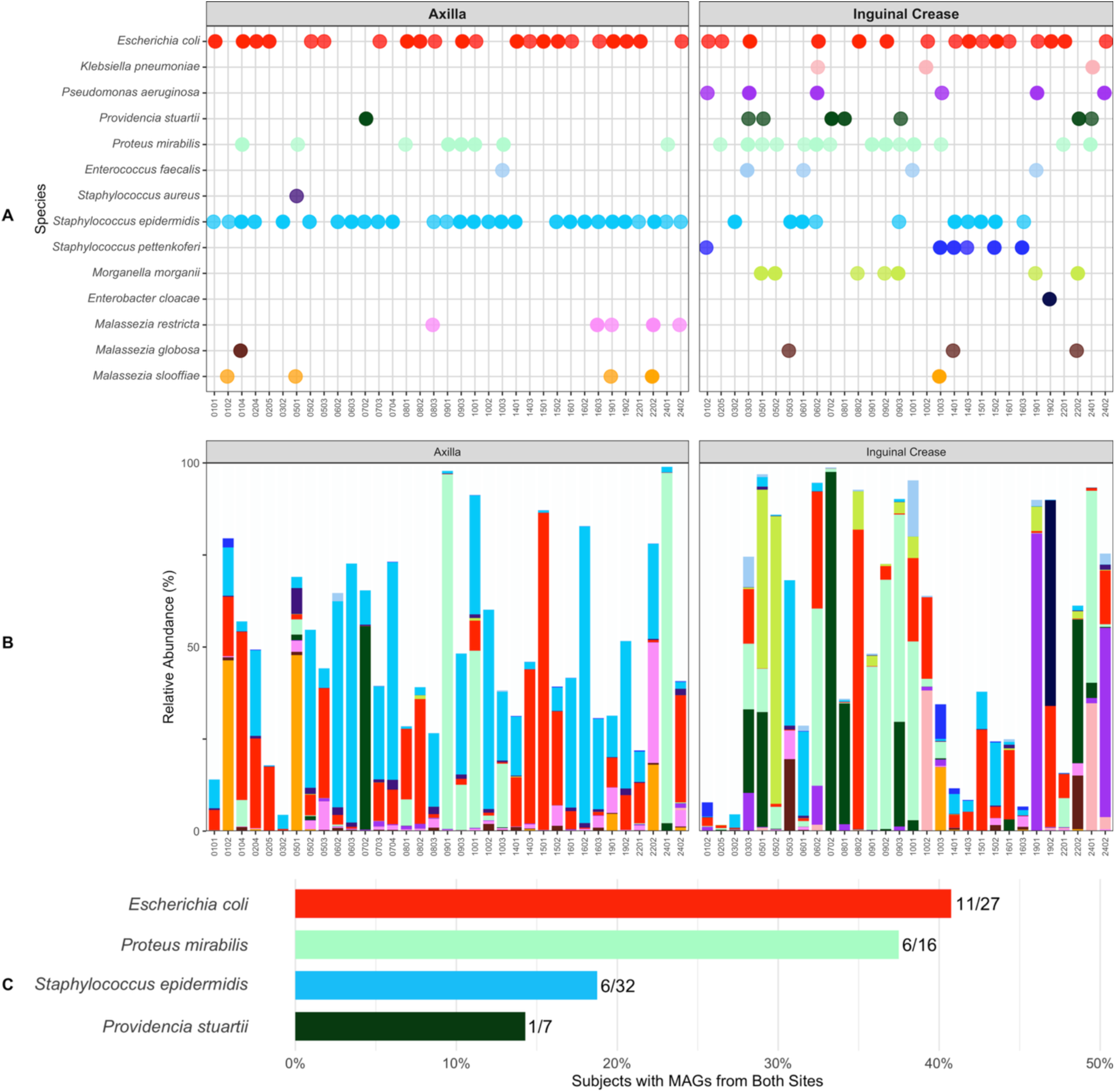
MDRO colonization of nursing home residents’ skin characterized by MAG recovery and abundance. (A) Recovery per resident at any timepoint of near-complete bacterial and fungal MAGs colored for representative species. Detailed overview of MAGs per resident per species are displayed in Supplementary Figure S2A. (B) Relative abundance of same representative bacteria and fungi in shotgun metagenomic sequencing samples. Colors are defined in panel A. (C) Frequency of MAG recovery from both Ic and Ax of the same resident.

Taken together, these findings point to the presence of persistent opportunistic pathogen populations on nursing-home residents’ skin rather than large compositional shifts over the short sampling period.

### MAG recovery reveals extensive skin colonization by opportunistic pathogens

From the skin shotgun metagenomic data generated from 207 samples, we recovered 1,185 MAGs, including 1,160 bacterial and 25 fungal bins, with a median of four MAGs per sample. Taxonomic classification revealed both skin commensals and clinically relevant microbes such as *Escherichia coli* (n=68 MAGs; 60 near-complete), *Staphylococcus epidermidis* (n=85; 77 near-complete), *Proteus mirabilis* (n=40; 34 near-complete), *Providencia stuartii* (n=17; 16 near-complete), and *Pseudomonas aeruginosa* (n=11; 10 near-complete) (Figure 2A; Supplementary Figure S2A). Fungal MAGs were largely *Malassezia* spp., consistent with typical skin mycobiota (Figure 2A; Supplementary Figure S2B). MAG abundances correlated with read-level detection across samples (Figure 2B), confirming analytic robustness.

Dual-site colonization of a resident was common for *E. coli*, with concurrent detection in axilla and inguinal crease in 11 of 27 colonized residents (40.7%), and for *P. mirabilis* in 6 of 16 residents (37.5%). *S. epidermidis* and *P. stuartii* displayed lower dual-site co-occurrence rates (18.8% and 14.3%, respectively) (Figure 2C). These findings suggest that high-burden colonizers can simultaneously occupy multiple skin niches, potentially increasing transmission risk within long-term care settings.

### Metagenomics reveals dominant MDR *E. coli* ST93 missed by culture

Genomic analyses further characterized the *E. coli* that colonized resident skin at the strain level. ESBL-selective culturing recovered *E. coli* from 4 of 38 residents, with genomic sequence analysis further delineating that 3 of these residents were colonized by *E. coli* ST131 and 1 resident was colonized by *E. coli* ST648. These four *E. coli* genomes carried multiple antibiotic resistance genes including *bla*_CTX-M-15_ and *bla*_OXA-1_ genes, which are known to confer an ESBL phenotype (Red asterisk in Figure 3A; Table S2). By contrast, MAG assembly from the skin metagenomic samples recovered *E. coli* MAGs from 27 residents, (Figure 3A, Table S2). *E. coli* ST131 MAGs were recovered from 4 residents, while half of all resident samples (19/38) yielded *E. coli* ST93 MAGs. None of these *E. coli* ST93 MAGs carried *bla*_CTX-M_ genes, nor was the *bla*_CTX-M_ gene detected in the corresponding skin metagenomes, which likely explains why these organisms did not grow on ESBL selective plates (Figure 3B, red box). However, the *E. coli* ST93 MAGs still carried a broad MDR repertoire such as *bla*_TEM,_ AmpC-family β-lactamases, aminoglycoside, fluoroquinolone, macrolide, sulfonamide, and tetracycline resistance genes (in the absence of other Proteobacteria on the contigs as confirmed by strainGE and BLASTn) (Figure 3B; Supplementary Figure S3). Moreover, an *E. coli* ST93 isolate was recovered in the wider PROTECT study from a California nursing home (not in our microbiome cohort) carrying *bla*_CTX-M-1_ on an *Inc*I1 plasmid, demonstrating the lineage’s genetic capacity to acquire ESBL determinants. Together, these data reveal a two-tiered colonization landscape: low-prevalence ESBL producing *E. coli* ST131 detected by cultivation and metagenomics and more prevalent MDR *E. coli* ST93 detected via metagenomics (Figures 3A-B, Table 2).

**Figure 3.**
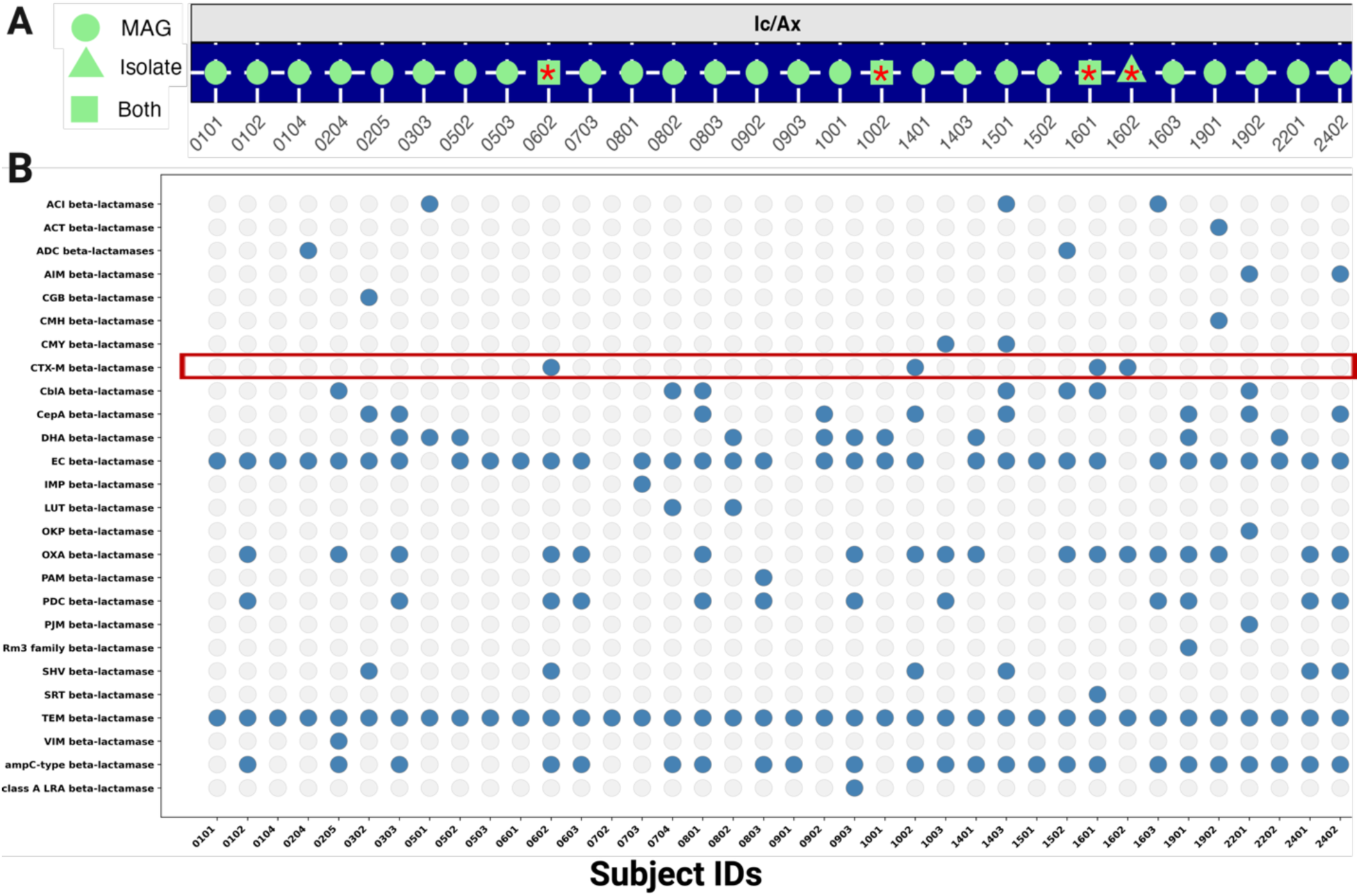
Nursing home residents’ skin colonization with multidrug resistant *E. coli*. (A) Detection of *E. coli* across residents with metagenome-assembled genomes (MAGs; circles), cultured isolates from ESBL-selective media (triangles), or both (squares). The red asterisk marks the β-lactamase gene content (i.e. carried *bla_CTX-M-15_* and *bla_OXA-1_*.). of cultured *E. coli* isolates from ESBL-positive residents. (B) Presence/absence matrix of β-lactamase gene families recovered from metagenomic sequencing across all residents. Detection of *bla_CTX-M_* genes (highlighted in red box) was confined to residents with cultured ESBL-producing isolates. In contrast, *E. coli* ST93 MAGs lacked *bla_CTX-M_* but encoded other β-lactamase families and multidrug resistance determinants (see Supplementary Figure S3).

### *E. coli* ST93 and ST131 persist on skin after bathing

Next, we leveraged our sampling on multiple days to assess whether *E. coli* strains identified by near-complete MAGs persisted on nursing home residents’ skin after bathing. Longitudinal sampling (two to three visits per resident) revealed stable *E. coli* colonization over 12-48 h post-bathing (Figure 4). Persistent detection post-bathing was observed in half the residents who were colonized by *E. coli*, including residents 0204, 0205, 0801, 0802, 1501, and 1502 carrying ST93, and 0802 and 0902 carrying ST131. Several of these residents had a documented bath between skin sampling, and the body site continued to be colonized with *E. coli* of the same sequence type. For example, *E. coli* ST93 was detected on the axilla and inguinal crease of resident 1502 at both 12 and 24 hours post-bath.

**Figure 4.**
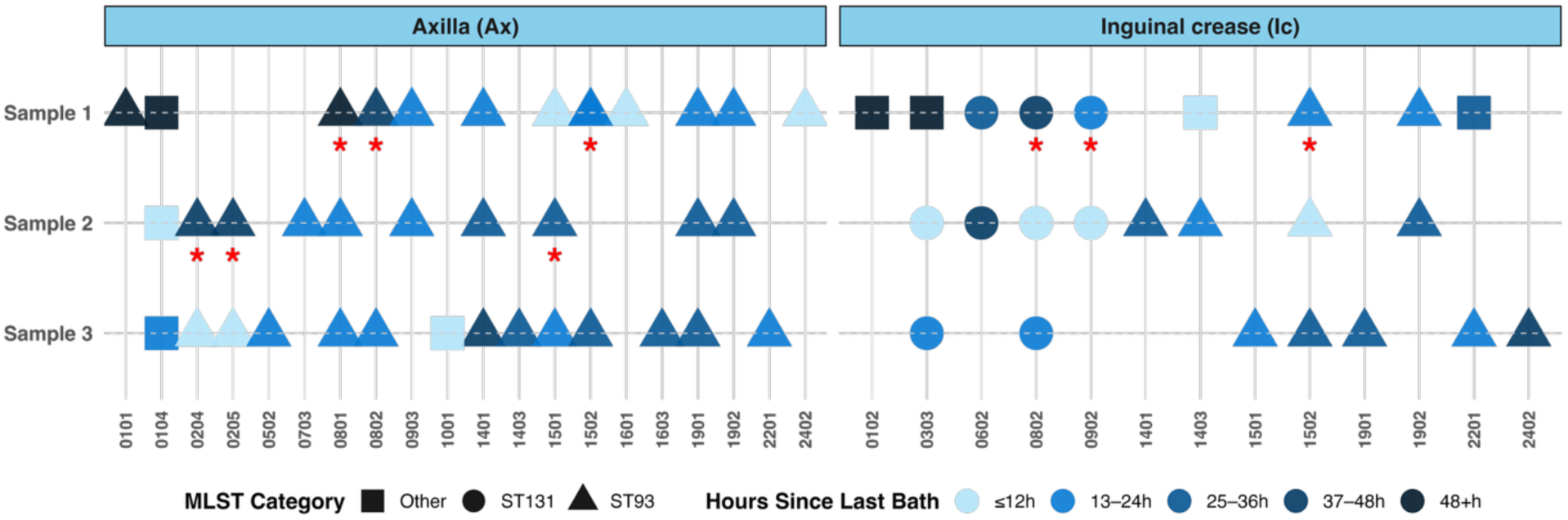
Persistent colonization of *E. coli* ST131 and ST93 in NH residents’ skin post-bathing. Reading from top to bottom, this figure shows persistent detection of near-complete *E. coli* ST131 and ST93 MAGs after bathing from the same resident at two skin sites. Shape represents STs, with the color indicating time since last bath ranging from 12h (lighter) to 48h (darker). The x-axis lists NH residents, and the y-axis indicates sampling events. Red asterisk (*) indicates when bathing occurred between sampling and is only applied for ST131 and ST93. Gaps represent no *E. coli* MAG detected.

### Regional clonal transmission of *E. coli* ST93 and diverse ST131 introductions

Consistent with the MLST typing, a combined phylogenetic analysis of *E. coli* MAGs and isolate genomes (Figures 5A-B) revealed two distinct clusters (ST93, ST131). A high-resolution SNP-based phylogeny of *E. coli* ST93 genomes showed a single clade circulating regionally across 15 NHs (Figure 5C), with ≤30 SNPs between any pair and ≤5 SNPs within the same individual (Figure 5D; Supplementary Table S5). Notably, 27 residents (71.1%) across 9 facilities shared clonally related *E. coli* ST93 strains (≤10 SNPs), suggesting widespread transmission or common-source exposure. The clonal, regional strains from this study were however >100 SNPs away from ESBL-producing *E. coli* ST93 obtained in another California NH in 2019, as well as other ST93 genomes from elsewhere in the country (Figure 5C).

**Figure 5.**
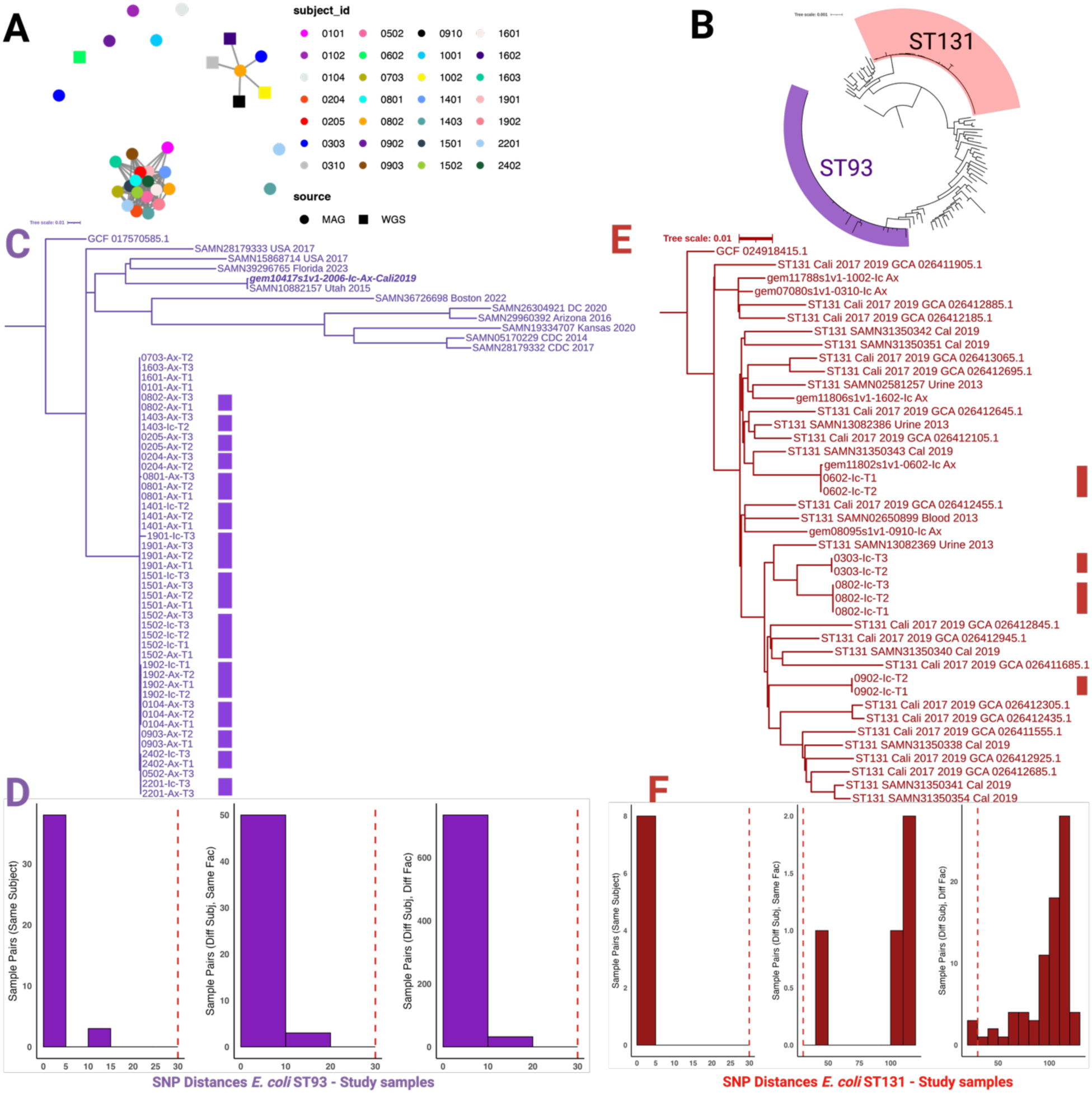
Comparative genomic analysis of *E. coli* strains from NH resident skin. (A) Network analysis of pairwise average nucleotide identity (ANI) among *E. coli* MAGs and isolate genomes from this study and the four isolate genomes from outside the cohort. Each node represents the highest-quality *E. coli* genome per resident, colored by resident. Circles denote MAGs and squares denote isolate genomes. Edges connect genomes with ≥99.95% ANI, indicating high genetic relatedness. (B) Phylogenetic tree based on single-copy marker genes, including *E. coli* isolate genomes, MAGs from this study, and public reference genomes. Clades corresponding to *E. coli* ST131 and ST93 are highlighted in red and purple, respectively. (C) Phylogenetic tree of *E. coli* ST93 genomes from this study alongside representative references, inferred from core SNPs. The purple blocks along the tree mark samples from the same resident. gem10417s1v1_2006_Ic-Ax is the ESBL-producing *E. coli* ST93 obtained in another California NH in 2019. MAGs are labeled with residentID_bodysite_timepoint; (D) Histogram of pairwise core genome SNP distances among study-derived *E. coli* ST93 genomes. The red dashed line marks a 30-SNP threshold for strain-level relatedness. (E) Phylogenetic tree of *E. coli* ST131 genomes from study samples and public references, inferred from core SNPs. The red blocks along the tree mark samples from the same resident. (F) Histogram of pairwise core genome SNP distances between study-derived *E. coli* ST131 genomes and public *E. coli* ST131 references. The red dashed line denotes a 30-SNP threshold used to infer potential strain sharing. Abbreviations: Diff Subj = different resident; Diff Fac = different facilities/NHs.

In contrast, *E. coli* ST131 genomes displayed greater diversity. While longitudinal samples within individuals were clonal (0-1 SNPs; Supplementary Table S6), intra-facility comparisons between residents typically exceeded 40 SNPs, suggesting multiple introductions. A few inter-facility comparisons (e.g., residents 0303 and 0802) approached potential transmission thresholds (28-52 SNPs) (Figures 5E-F). Comparisons with public genomes from California highlighted occasional proximity to clinical *E. coli* ST131 isolates from California, implying regional overlap but overall high diversity. These results revealed a dominant, clonally conserved *E. coli* ST93 lineage spreading widely, contrasted with sporadic, diverse *E. coli* ST131 introductions.

### Methicillin-resistant *S. epidermidis* ST2 persists on skin and spreads across facilities

To assess how *E. coli* dynamics within and across NHs compared to other high prevalence MDROs, we next explored the persistent post-bathing detection and sharing of methicillin resistant *Staphylococcus epidermidis* (MRSE) ST2 in nursing home residents. *S. epidermidis* was detected in 84% of residents, with ST2 MRSE MAGs found in 14 residents (36.8%) (Figure 6A). The *mec*A gene was detected in 23 of 28 *S. epidermidis* ST2 MAGs as well as in the remaining 5 samples co-colonized with *S. aureus*. Persistent MRSE colonization was observed over 12-48 hours post-bathing in residents 1001, 1002, 0104, 0601, and 0602 (Figure 6B; Supplementary Figures S4-S5, Table S7). Within-host SNP distances ranged from 0-3, indicating clonal persistence. ANI and SNP-based networks revealed closely related *S. epidermidis* ST2 strains circulating within and across facilities (Figures 6C-E). Among the many examples, highly related strains (5-9 SNPs) were shared by residents 0601 and 0602 in NH 06, as well as related strains (18 SNPs) in residents 1001 and 1002 in NH 10. Strains from different NHs were also highly related (e.g., 0104 and 0602 shared strains with only 5 SNPs difference; 0704 and 0602 with 10 SNPs; 0104 and 0704 with 10 SNPs), suggesting regional dissemination of a predominant MRSE ST2 clone persisting on skin and potentially spreading between nursing homes.

**Figure 6.**
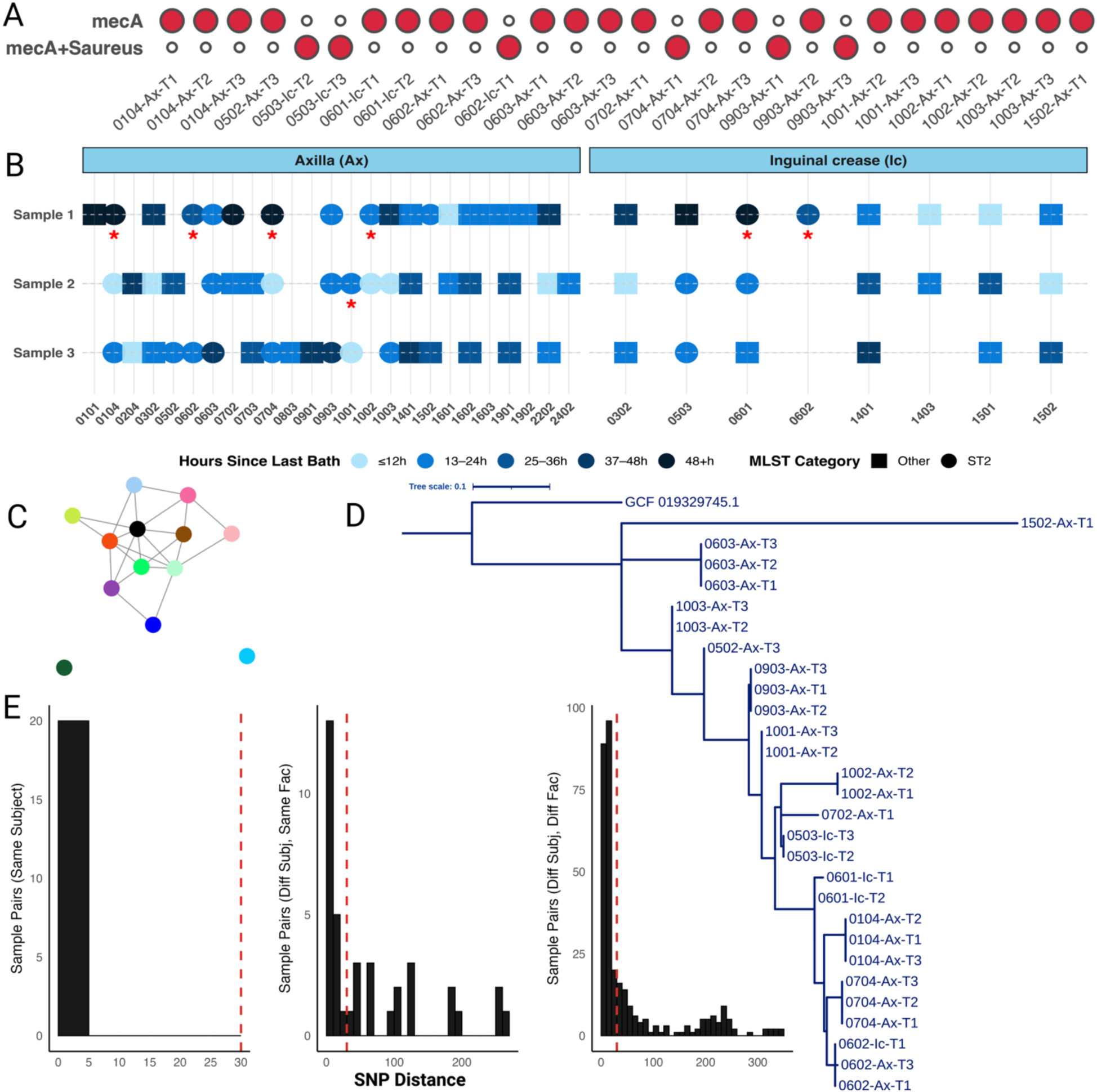
Persistence and resident strain sharing of methicillin resistant *Staphylococcus epidermidis* ST2 within and between nursing homes. (A) Detection of the *mecA* gene across metagenomic samples. Each circle represents a metagenomic sample. Red-filled circles indicate detection of *mecA*; *mecA*+Saureus is indicative of co-presence by *S. aureus* in the metagenomic reads as identified by StrainGE. (B) Persistent recovery of *S. epidermidis* ST2 MAGs before and after bathing, stratified by body site (axilla [Ax] and inguinal crease [Ic]) and organized by resident. Each tile represents a sample; colored circles indicate recovery of an *S. epidermidis* ST2 MAG and are colored by time since last bath. Red asterisks mark persistent detection of *S. epidermidis* ST2 across multiple timepoints in the same resident. In the facet “Inguinal Crease” for resident 0602 there is an asterix but no isolate indicated for Sample 2 or 3 because the individual had a bath and a subsequent MAG but this MAG did not meet the near complete criterion. (C) Average nucleotide identity (ANI) network of *S. epidermidis* ST2 MAGs. Nodes represent individual *S. epidermidis* ST2 genomes, colored by resident, and edges connect genomes sharing ≥99.95% ANI. The clustering of genomes from different residents and facilities reflects the circulation of a highly conserved *S. epidermidis* ST2 lineage across the nursing homes. (D) Maximum-likelihood phylogenetic tree of *S. epidermidis* ST2 rooted to the *S. epidermidis* type strain (GCF_001922515.1). (E) Histogram of pairwise SNP-based genomic distance between *S. epidermidis* ST2 samples. Red dashed lines indicate the 30 SNP threshold for defining closely related strains.

### Proteus, Enterococcus and Providencia exhibit persistence and facility-level spread

We broadened our analyses of strain transmission dynamics to a range of other, less prevalent, microbes in nursing home residents. *Proteus mirabilis* was detected in 16 residents (42.1%), including three ESBL-producing isolates carrying *bla*_CTX-M-15_ (Figure 7A-B; Table 2). *E. faecalis* was detected in 7 residents (18.4%), with VRE isolates typed as ST109 and ST778 (Table 2).

**Figure 7.**
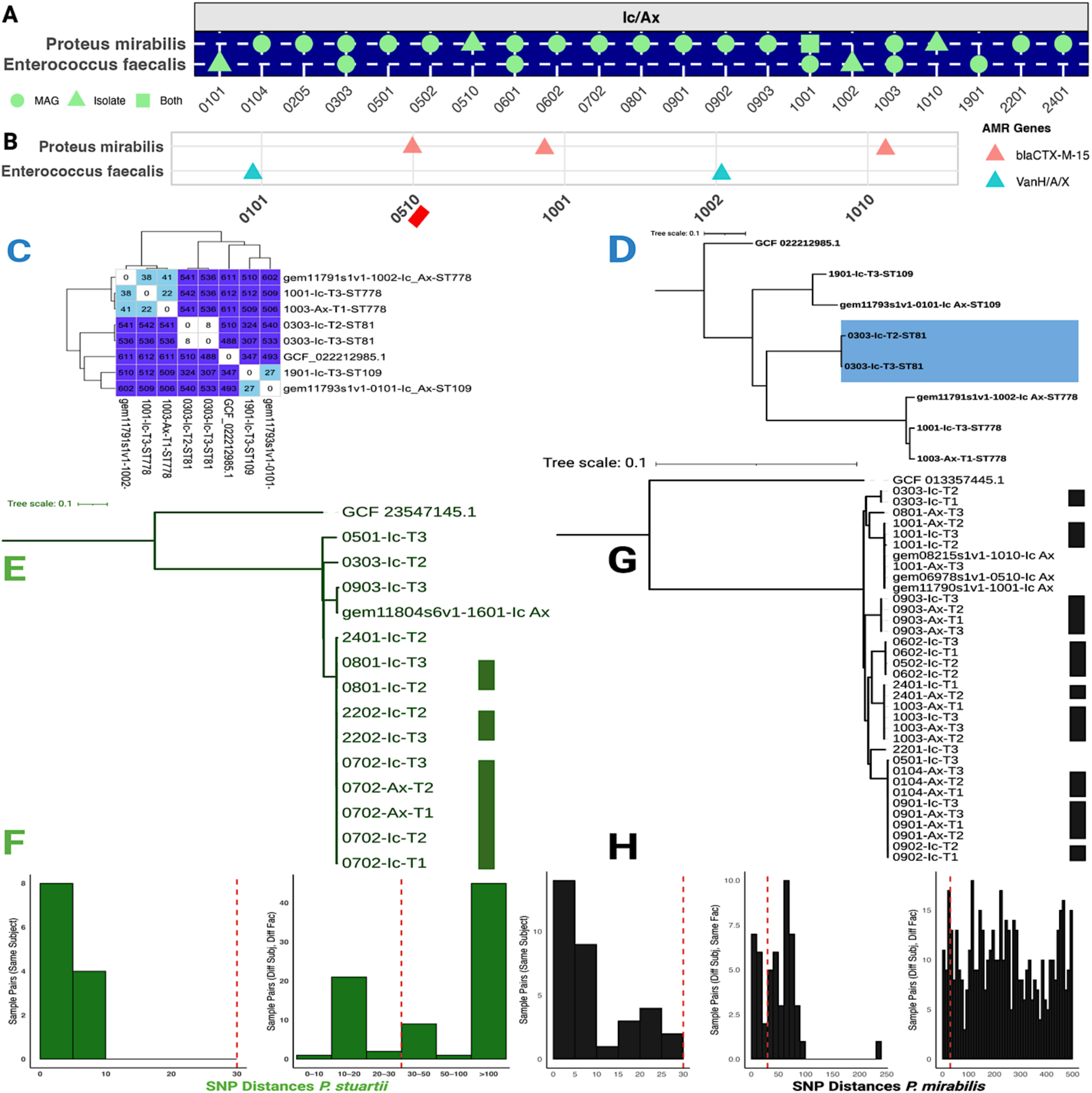
Genomic relatedness of healthcare-associated microbes colonizing nursing home residents. (A) Detection of *Proteus mirabilis* and *Enterococcus faecalis* on skin across NH residents (n = 28), based on recovery as metagenome-assembled genomes (MAGs), isolates, or both. (B) Antimicrobial resistance gene detection in isolate genomes. *bla_CTX-M-15_* was found in *P. mirabilis* isolates, while *vanHAX* operon genes (*vanH*, *vanA*, *vanX*) were identified in *E. faecalis*. The red black below resident 0510 is to indicate that it is one of the external resident to our cohort but coming from the same NHs as indicated in the methods. (C) Pairwise SNP distance heatmap among *E. faecalis* strains. Lower values indicate greater genomic similarity. Samples starting with the identifier “gem” are isolate genomes and helps to differentiate from MAGs. (D) Maximum likelihood phylogeny of *E. faecalis* isolates and MAGs, with clustering by resident and timepoint. (E) Phylogenetic tree of *Providencia stuartii* MAGs shows resident-specific clustering. (F) SNP histogram for *P. stuartii*. (G) Phylogenetic tree of *P. mirabilis* strains. (H) SNP histogram for *P. mirabilis*.

Both species displayed within-host persistence and inter-resident sharing. For *P. mirabilis*, SNP distances of 0-7 were observed within facilities and between facilities (e.g., 1001-Ax-T3 identical to 0903-Ax-T2) (Figures 7G-H; Supplementary Table S9). *E. faecalis* isolates from 1001-1003 clustered phylogenetically (22-41 SNPs), suggesting facility-associated circulation, while inter-facility relatedness (1901 vs. 0101, 27 SNPs) pointed to a possible regional reservoir (Figures 7C-D; Supplementary Table S8).

*Providencia stuartii* was detected in 7 residents (18.4%), showing clonal persistence within individuals and near-identical strains (≤13 SNPs) shared between facilities (Figure 7E-F). Additional MDROs included *P. aeruginosa* and *M. morganii* (Supplementary Figure S6), often showing within-host clonality and occasional inter-resident sharing (<30 SNPs), while *K. pneumoniae* (ST20, ST1842) and *S. pettenkoferi* colonization remained largely resident-specific (Supplementary Figures S7-S8).

### Polymicrobial strain sharing reveals complex transmission networks

Finally, we aggregated our findings to explore possible polymicrobial strain sharing among nursing home residents. Five resident pairs harbored clonally related strains from ≥2 MDRO species, underscoring complex transmission networks. For example, residents 0902 and 0903 shared near-identical *P. mirabilis* (3-4 SNPs) and *E. coli* ST93 (1-3 SNPs) in NH 09 (Figure 7). Cross-facility polymicrobial sharing was observed between NHs 08 and 22 (*P. stuartii*, *M. morganii*, ≤30 SNPs). Overall, 27 residents across 9 NHs (71.1%) shared at least one clonally related MDRO strain with another resident (≤10 SNPs), highlighting extensive intra- and inter-facility dissemination of multidrug-resistant skin colonizers.

## Discussion

Antimicrobial resistance represents one of the greatest challenges to global health, with its burden projected to rise substantially in coming decades. Surveillance and response strategies have largely centered on acute-care hospitals and the livestock sector, leaving long-term care facilities, particularly nursing homes, underappreciated despite their role as persistent MDRO reservoirs^5,9^. Using a combined approach of shotgun metagenomics, metagenome-assembled genomes, and isolate genome sequencing, we characterized MDRO colonization on nursing home residents’ skin and revealed ecological persistence, regional dissemination, and multi-species sharing events that are largely invisible to current culture- and peri-rectal based diagnostics.

Our data demonstrate that MDRO colonization on skin is persistent rather than transient. Across multiple timepoints, clonal strains of *E. coli* (ST93, and ESBL-producing ST131), methicillin-resistant *S. epidermidis* (MRSE ST2), and vancomycin-resistant *E. faecalis* (ST109, ST778) persisted despite bathing intervals of 12-36 hours, consistent with previous findings that skin acts as a long-term reservoir for clinically relevant microbes capable of causing opportunistic infections^7^. Such persistence has implications for autoinoculation of wounds or medical devices, increased risk of infection, and ongoing environmental contamination that may sustain transmission cycles in NH settings.

One of the most striking findings was the widespread presence of *E. coli* ST93, detected in 71% of residents via MAGs yet absent from ESBL-selective culture plates. These strains lacked *bla*_CTX-M_ genes but carried a broad resistome, including *bla*_TEM_, AmpC β-lactamases, and resistance determinants for aminoglycosides, sulfonamides, and fluoroquinolones. The detection of an *E. coli* ST93 isolate in a neighboring nursing home carrying *bla*_CTX-M-1_ on a conjugative *Inc*I1 plasmid bordered by IS1380 elements provides direct evidence of this lineage’s potential to acquire ESBL genes, a configuration well documented in past ESBL epidemics. ^14,15^ Public data reveal additional high-risk variants of *E. coli* ST93, including isolates with *bla*_CTX-M-55_ and *bla*_NDM-1_ (SAMN28179333, SAMN39296765), as well as colistin resistance gene *mcr*-1 in Asia^16^, underscoring the lineage’s evolutionary potential to become a clinically significant MDRO. These results highlight that current resistance-targeted culturing may underestimate *E. coli* reservoirs in long-term care facilities, leaving silent dissemination unchecked.

Similarly, *S. epidermidis* ST2, a healthcare-adapted clone linked to device-associated infections in immunocompromised hosts^17,18^ was detected in 37% of residents, often shared clonally within and across facilities, with *S. epidermidis* ST2 MAGs differing by ≤10 SNPs. These strains frequently carried *mec*A and circulated regionally, yet none were captured by culturing since methicillin-resistant *S. epidermidis* was not specifically sought-after during culturing. These findings suggest that dominant, clinically relevant lineages may be silently spreading within NHs, escaping detection by routine clinical workflows focused on hospital pathogens that rarely involve skin sampling.

Our metagenomic analysis further demonstrated that nursing homes act as interconnected nodes in regional transmission networks rather than isolated microbial ecosystems. Clonally related *E. coli* ST93, *E. faecalis* ST109/ST778, *Proteus mirabilis*, and *Providencia stuartii* strains were shared both within and between nursing homes, consistent with recent genomic epidemiology studies reporting MDRO dissemination among NH residents^9, 19, 20^. Frequent resident transfers between NHs and hospitals^5^, combined with limited infection prevention resources and delayed implementation of mandatory programs^21^, may contribute to this silent, inter-facility spread. Our data suggest that colonized residents, even in the absence of active infection, may serve as long-term, mobile reservoirs bridging care settings and contributing to regional MDRO circulation.

In several cases, we identified polymicrobial strain sharing where resident pairs carried clonally related strains from two or more distinct MDRO species. These events occurred both within facilities and across different nursing homes, involving combinations of *E. coli* ST93, *S. epidermidis* ST2, *E. faecalis*, *Providencia stuartii*, and *P. mirabilis*. These polymicrobial colonization patterns highlight that transmission dynamics in nursing homes are not pathogen-specific but involve broader microbial consortia that may co-evolve, persist, and potentially exchange resistance genes on skin. Infection control strategies focused on single priority pathogens may therefore fail to interrupt the true ecological networks sustaining AMR in long-term care facilities. While our study did not integrate environmental samples like was done in other studies^8^, the use of shotgun metagenomics here gives a much wider overview on the constellation of culturable and non-culturable MDROs in the nursing homes.

Shotgun metagenomics provided critical insights into the skin resistome, detecting ARGs spanning multiple antibiotic classes, including β-lactams (*bla*_TEM_, *bla*_CTX-M_, *bla*_OXA_, *mec*A), aminoglycosides, sulfonamides, fluoroquinolones, phenicols, macrolides, and tetracyclines. Many of these genes were detected without corresponding culture isolates, highlighting the limitations of diagnostics dependent on phenotypic growth under selective conditions. For example, *E. coli* ST93 MAGs lacked ESBL determinants yet encoded multiple β-lactamases, illustrating a latent capacity for resistance that would remain undetected in standard screening. Although current binning algorithms limit direct plasmid-to-host ARG attribution^22,23^, read-level mapping confirmed key resistance determinants in high-risk lineages such as *E. coli* ST131 and ST93 (in the absence of other Proteobacteria on the contigs as confirmed by strainGE and BLASTn), supporting their potential clinical significance.

Collectively, the ecological and genomic data indicate that resistance-targeted culturing alone underestimates MDRO burden on skin. Integrating metagenomic surveillance alongside targeted culturing could close detection gaps and guide more effective infection-prevention strategies across long-term care and the healthcare networks they connect to.

This study has limitations. We sampled a maximum of three residents per nursing home, preventing detailed analysis of within-facility networks, including between roommates. Environmental sampling was not performed, restricting source tracking. Also, SNP-based thresholds (<30 SNPs) align with genomic epidemiology standards^9^ but cannot establish transmission directionality without denser temporal and epidemiological data.

Despite these limitations, our findings establish nursing home resident skin as a stable and overlooked reservoir of MDROs with potential for persistence and dissemination within and across facilities. The recurrent detection of polymicrobial strain sharing underscores that long-term care residents are exposed to overlapping reservoirs and transmission routes involving multiple resistant taxa. Culturing confirmed that these MDROs were viable and replicating on the skin while metagenomics enabled detection of additional pathogens of concern that were missed by selective culturing. Expanding surveillance frameworks to include skin as relevant site and nursing homes, employing genome-informed, multi-species strategies that integrate metagenomics with traditional culturing, could close critical detection gaps and guide more effective infection control. As the population of frail, immunocompromised individuals in long-term care grows, addressing colonization and transmissions within nursing homes will be crucial to curb the spread of antimicrobial resistance within healthcare ecosystems.

## Methods

### Study design, participants and skin sample collection

This skin microbiome study was nested within Project PROTECT (NCT03118232), a cluster-randomized trial of 28 nursing homes in California, USA. ^5^ Between September and December 2018, a convenience sample of residents from facilities assigned to both routine care and decolonization arms were enrolled. A total of 38 residents from 15 NH were enrolled and contributed usable samples (Table S1). Two skin sites (inguinal crease and axilla) were sampled at 2-3 visits per resident between 12h and 48h after bathing (Figure 1A). In total we analyzed 223 samples including 207 skin swabs, 6 sample controls (air swabs at sampling site), 6 negative controls (reagent-only controls), and 4 positive controls (Mock Community standard ZymoD6306, ZymoBIOMICS, Zymo Research, California, USA). The study was approved by the Institutional Review Board at the University of California, Irvine, and informed consent was obtained from all participants or their representatives.

### Culture-based microbiology

All 207 skin swabs and the 6 air swabs were plated on CHROMagar™ ESBL, CHROMagar™ VRE and CHROMagar™ MRSA to selectively isolate ESBL-producing Enterobacterales, vancomycin-resistant *Enterococcus* (VRE) and methicillin-Resistant *Staphylococcus aureus* (MRSA). Colonies were sub-cultured, and DNA extraction was performed using the QIAmp 96 DNA QIAcube HT kit or the Omega Bio-Tek Universal Pathogen Core Kit with modifications. Library preparation used Nextera XT kits, and sequencing (of 23 isolates excluding MRSA isolates) was performed on Ilumina NovaSeq X+ (Table S2). Sequence reads were assembled with SPAdes^24^ v3.14.1, and quality assessed using QUAST^25^ v5.2.0 and CheckM2^26^ v1.0.2. Isolate genomes were analyzed for antimicrobial resistance genes with Abricate (using ResFinder v4.0 database^27^).

### Skin shotgun metagenomic sequencing and analysis

DNA from 207 skin swabs, 6 negative air swabs, 6 reagent-only controls, and 4 samples of the mock microbial community standard was extracted as previously described. ^10^ Nextera libraries were prepared and shotgun metagenomic sequencing was performed as paired-end reads (151 bp) on Illumina NovaSeq6000. Shotgun metagenomic reads were trimmed of adapters and low-quality bases with Cutadapt v4.0, and mapped to the T2T-CHM13v2.0 human reference genome using Bowtie2 v2.4.5 to remove host reads. ^28^ Quality metrics were assessed with FastQC and MultiQC.

Metagenome-assembled genomes (MAGs) were reconstructed using metaSPAdes v3.15.0 and MetaWRAP v1.2.2^29^, integrating MetaBAT2, MaxBin2, and CONCOCT for binning. MAGs were retained if completeness ≥50% and contamination ≤5% (CheckM2 ^26^v1.0.2) as previously defined^10^. However, for strain sharing and persistence analyses near-complete MAGs were used (Median completeness of 97.45% and a median contamination of 0.45%). Chimeric assemblies were identified with GUNC v1.0.5^30^ and removed. Taxonomic assignment used GTDB-Tk (release 207^31^). Eukaryotic MAGs were screened with EukCC v2.1.1^32^ and classified using Mash v2.3^33^ and Mummer v3.23^34^ against fungal genome references (after downloading all fungal genomes in RefSeq on September 25^th^ 2024). MAG assembly statistics, gene content metrics, and quality scores are provided in Table S3.

Taxonomic composition of skin metagenomic samples was determined using Kraken2 v2.1.2^35^ with a custom database of the standard Refseq database (Refseq-Release226) enriched with dereplicated MAGs from this study. Abundance estimates were generated with Bracken v2.8.0 and imported into R (phyloseq v1.38). Antibiotic resistance genes (ARGs) were detected by metagenomic reads-mapping with RGI v5.2.1 using the CARD database v3.1.2. ^36^ Genes were considered present with ≥40% allele coverage and ≥70% nucleotide identity. In downstream analysis, ARGs were considered for *E. coli* when other Proteobacteria were absent on the contigs as confirmed by strainGE^37^ and BLASTn. Methicillin resistance was extrapolated by detection of *mecA* in *Staphylococcus epidermidis* MAGs.

### Strain-level analyses

Strain level analysis characterized MAGs and bacterial isolate genomes from this study as well as dereplicated public genomes from RefSeq database for each species of interest downloaded on September 2^nd^, 2024. The genomic characterization of the ESBL Enterobacterales and VRE isolates recovered from NH residents and four comparative genomes from additional residents from the same facilities as part of the wider PROTECT trial^5^ are provided in Table S2.

We performed Multi-locus Sequence Typing (MLST) for lineage assignment with mlst v2.23.0, using default settings and published pubMLST schema (https://pubmlst.org/, MLST Achtman). Phylogenomic analyses based on single-copy marker genes was built with GToTree^38^ and core-genome SNP based phylogenetic analysis for maximum likelihood trees with RaxML ^39^ following previously described procedures^9^. Pairwise average nucleotide identity (ANI) was computed using FastANI v1.34.1^40^ to infer genomic similarity thresholds. Core-genome SNP profiling was performed with Snippy v4.4.1, with recombination masking using Gubbins v2.4.1^41^. SNP distances were calculated with snp-dists v0.8.2. Read-based strain detection from metagenomic sequences was performed using StrainGE v1.3.3^37^ with a database constructed with the combination of isolate genomes and dereplicated public genomes. A threshold of 30 SNPs was considered for phylogenetic relatedness based in the targeted species in NH settings from previous studies^9^.

### Statistical analyses

All downstream analyses were performed in R v4.3.2. Taxa present at <0.1% relative abundance or in <5% of samples were dropped. Alpha and beta diversity metrics were computed and differences across body sites and time points were assessed by PERMANOVA. Pairwise SNP distances and phylogenetic relationships were visualized using ggplot2 and iTOL.

## Data Availability

Complete data availability information, including SRA accession numbers for metagenomic and isolate reads, are provided in Supplementary Tables 1 and 2, respectively. The shotgun metagenomic reads are available in SRA under the BioProject ID: PRJNA1330355, while the sequence reads for isolate genomes were submitted to SRA under the BioProject PRJNA1301865.

Full computational workflows, detailed software parameters, command-line scripts and codes are provided on github.com/skinmicrobiome.

## Acknowledgements

We thank the nursing home residents and the staff at the facilities that participated in the study. The contributions of the NIH authors are considered Works of the United States Government. The findings and conclusions presented in this paper are those of the author(s) and do not necessarily reflect the views of the NIH or the U.S. Department of Health and Human Services.

We also thank the African Postdoctoral Training Initiative program funded by the Fogarty International Center (NIH) and the Gates Foundation under the African Academy of Sciences through which Dr Hounmanou was employed at the NIH.

## Author contributions

JAS, SC, GMG, RDS, HHK and SSH conceived and designed the study; GMG, RDS, CD and SSH oversaw the collection and processing of samples and culturing; YMGH, SC, DMP, MT, AME, CJW analyzed and interpretated genomic data and conducted bioinformatic analysis; YMGH, GMG, SSH and JAS wrote the paper with input from all authors. The authors read and approved the final manuscript.

## Supplemental material

### Supplementary Tables

SUPPLEMENTARY Table 1: Shotgun metagenomic data mapping file describing samples

SUPPLEMENTARY Table 2: Isolate genome sample and sequence characteristics

SUPPLEMENTARY Table 3: MAGs recovered with taxonomic and quality information

SUPPLEMENTARY Table 4: Antibiotic resistance gene mapping of shotgun metagenomic samples

SUPPLEMENTARY Table 5: SNP analysis for *E coli* ST93 MAGs and reference genomes

SUPPLEMENTARY Table 6: SNP analysis for *E coli* ST131 isolates and MAGs

SUPPLEMENTARY Table 7: SNP analysis for *S epidermidis* ST2 MAGs

SUPPLEMENTARY Table 8: SNP analysis for *Providencia stuartii* isolates and MAGs

SUPPLEMENTARY Table 9: SNP analysis for *Proteus mirabilis* isolates and MAGs

### Supplementary Figures

**Supplementary Figure S1:**
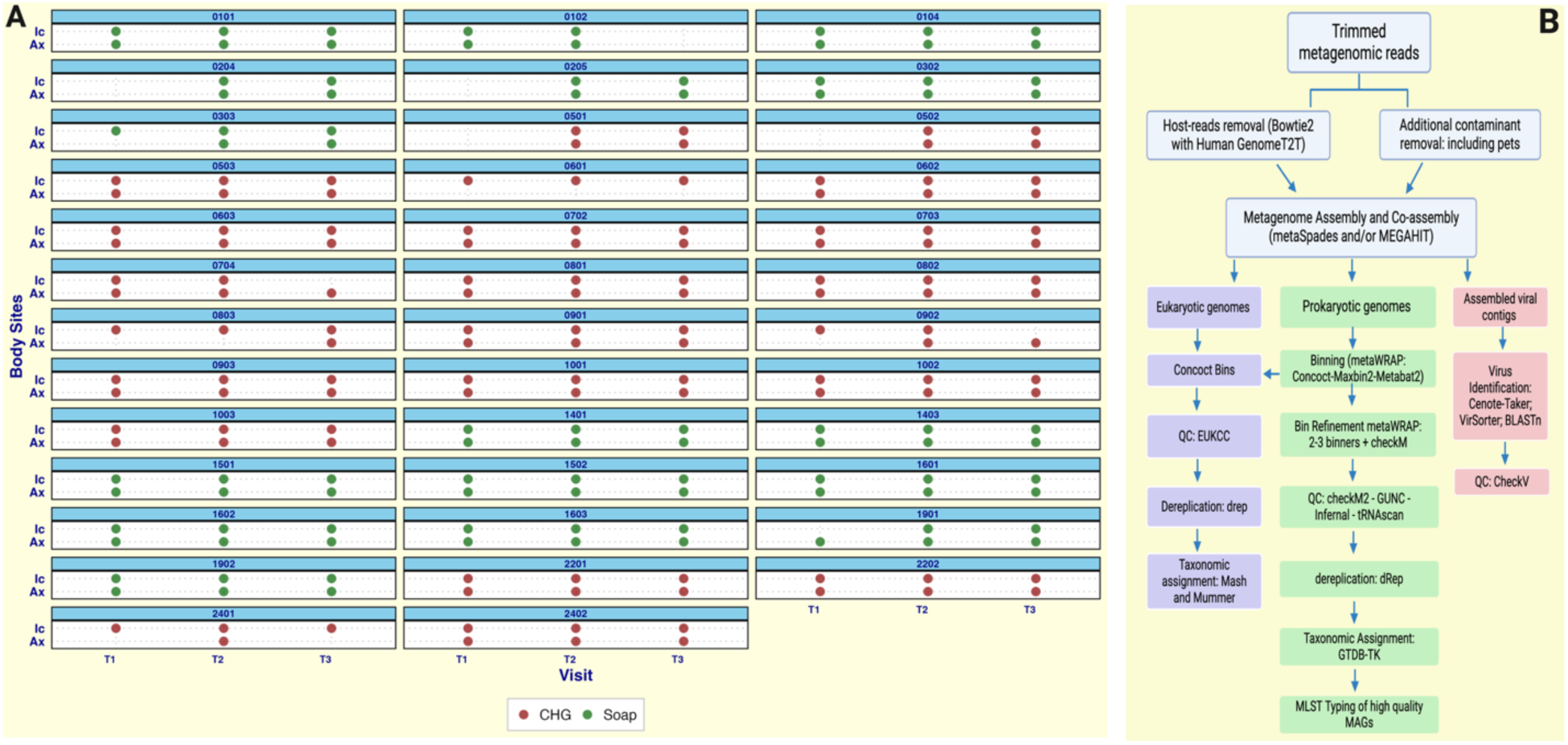
**(A)** Overview of samples analyzed in this study and (B) overview of the analytical pipeline from sequences to MAGs. CHG = NHs where bathing is done with chlorhexidine gluconate. Soap = NHs where bathing is done with regular soap.

**Supplementary Figure S2:**
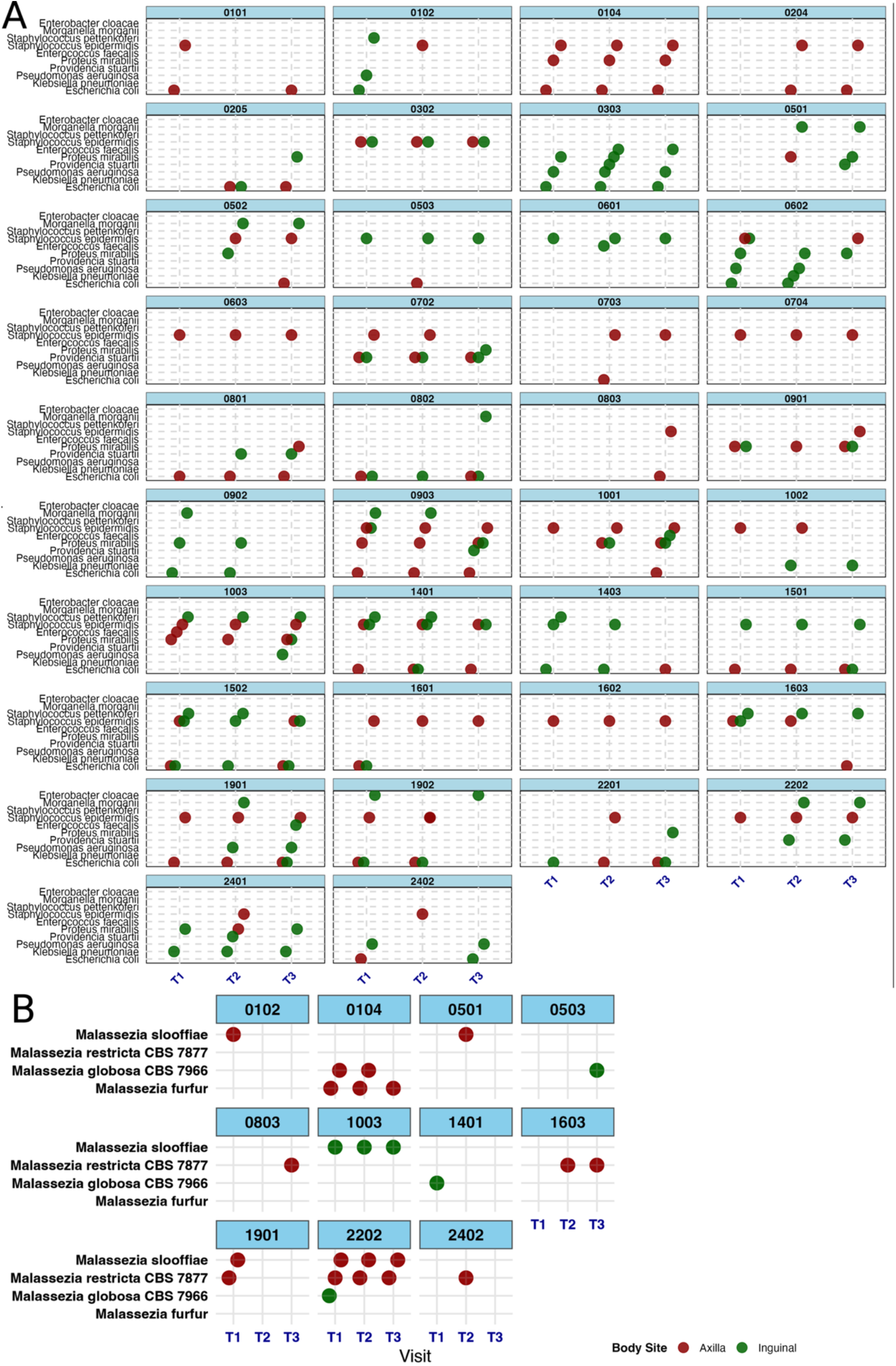
Comprehensive overview of metagenome-assembled genomes (MAGs) from selected bacterial (A) and fungal (B) species of clinical relevance analyzed in this study.

**Figure S3.**
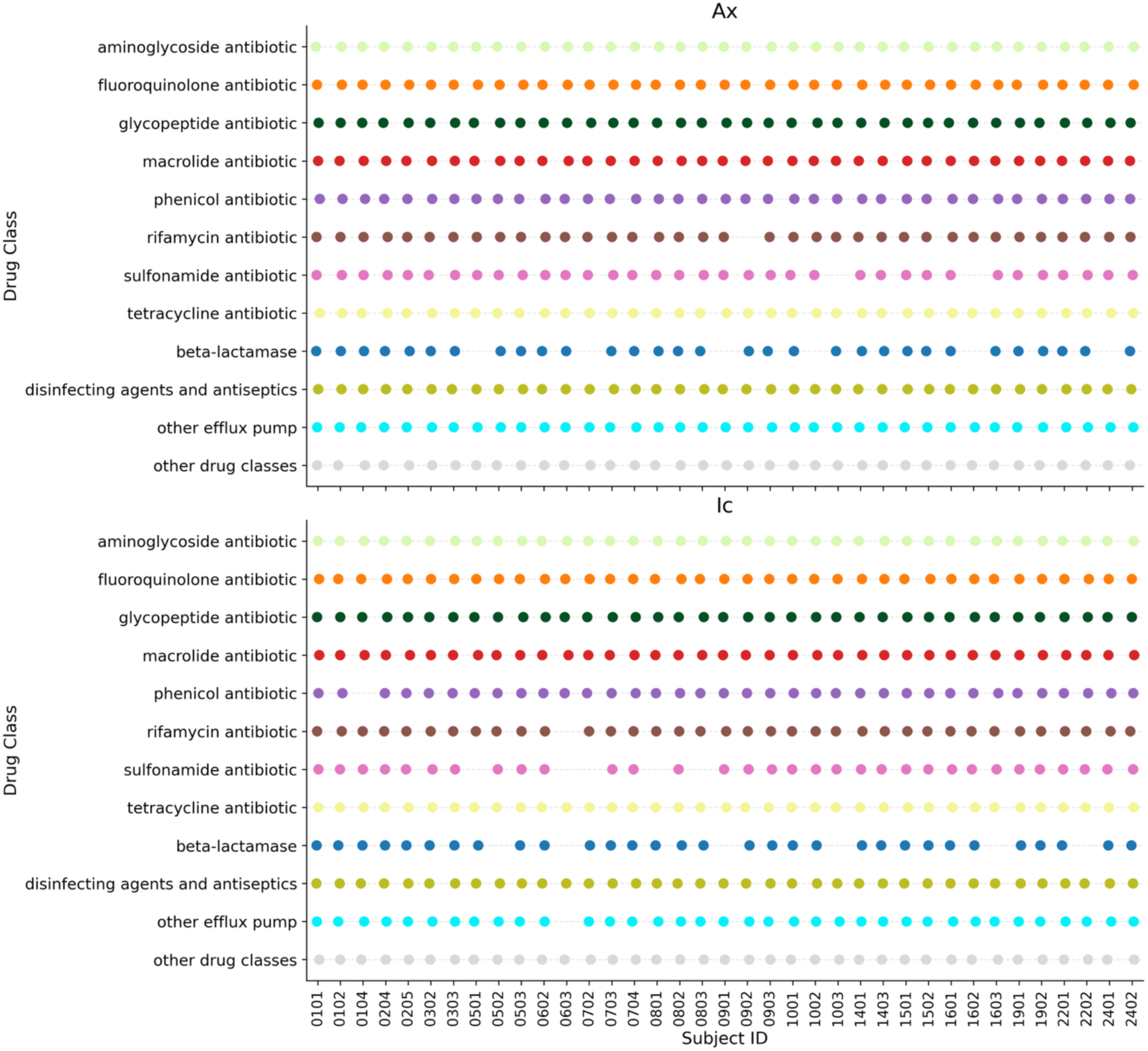
Antibiotic resistance classes by resident across body sites. Dot plot showing the presence (colored dots) or absence (blank space) of major drug classes across individual residents, stratified by body site: axilla (Ax) and inguinal crease (Ic). Each row corresponds to a drug class, and each column to a resident ID.

**Figure S4:**
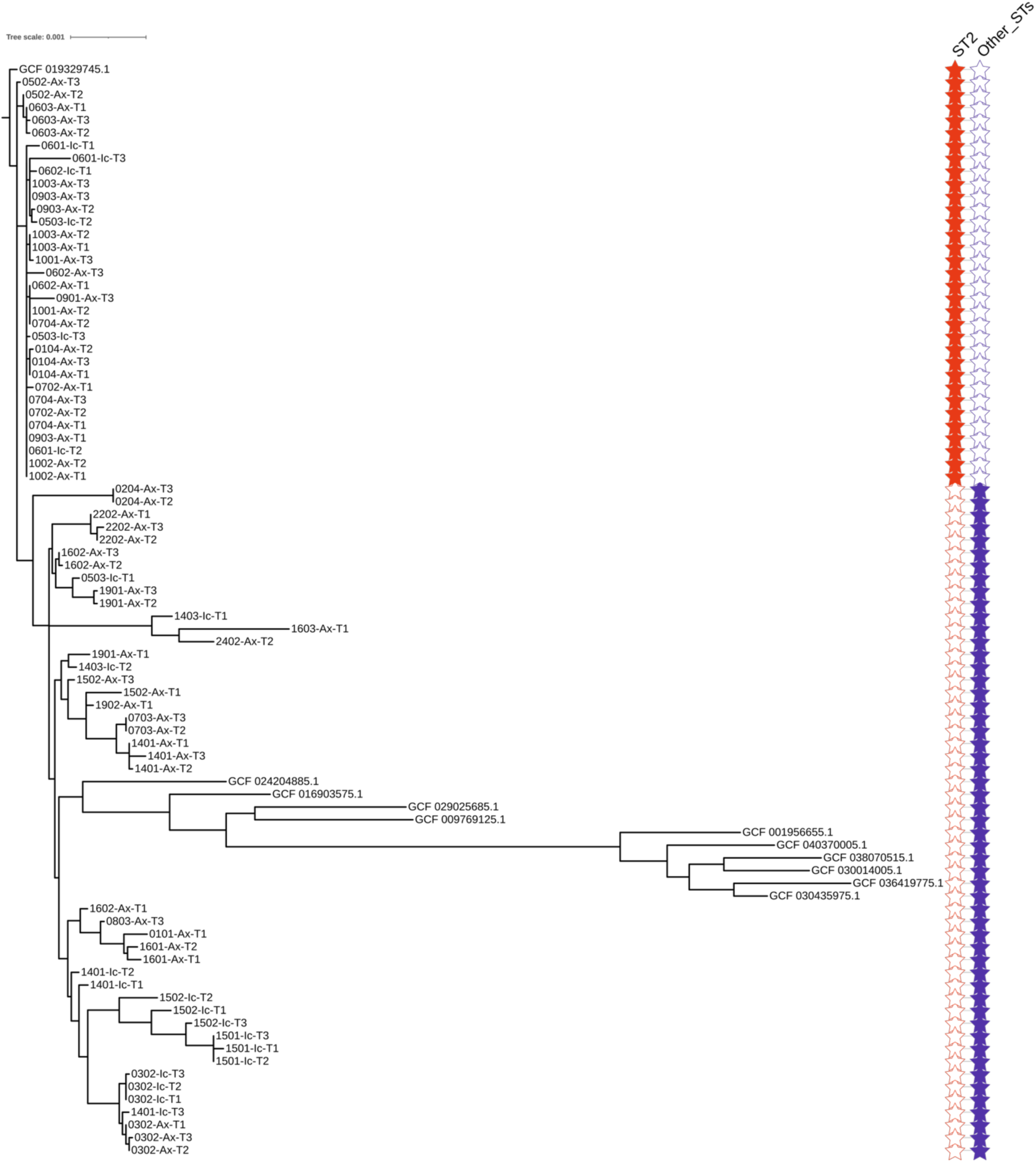
Overview of single copy marker gene tree of all *Staphylococcus epidermidis* analyzed along with public genomes

**Figure S5.**
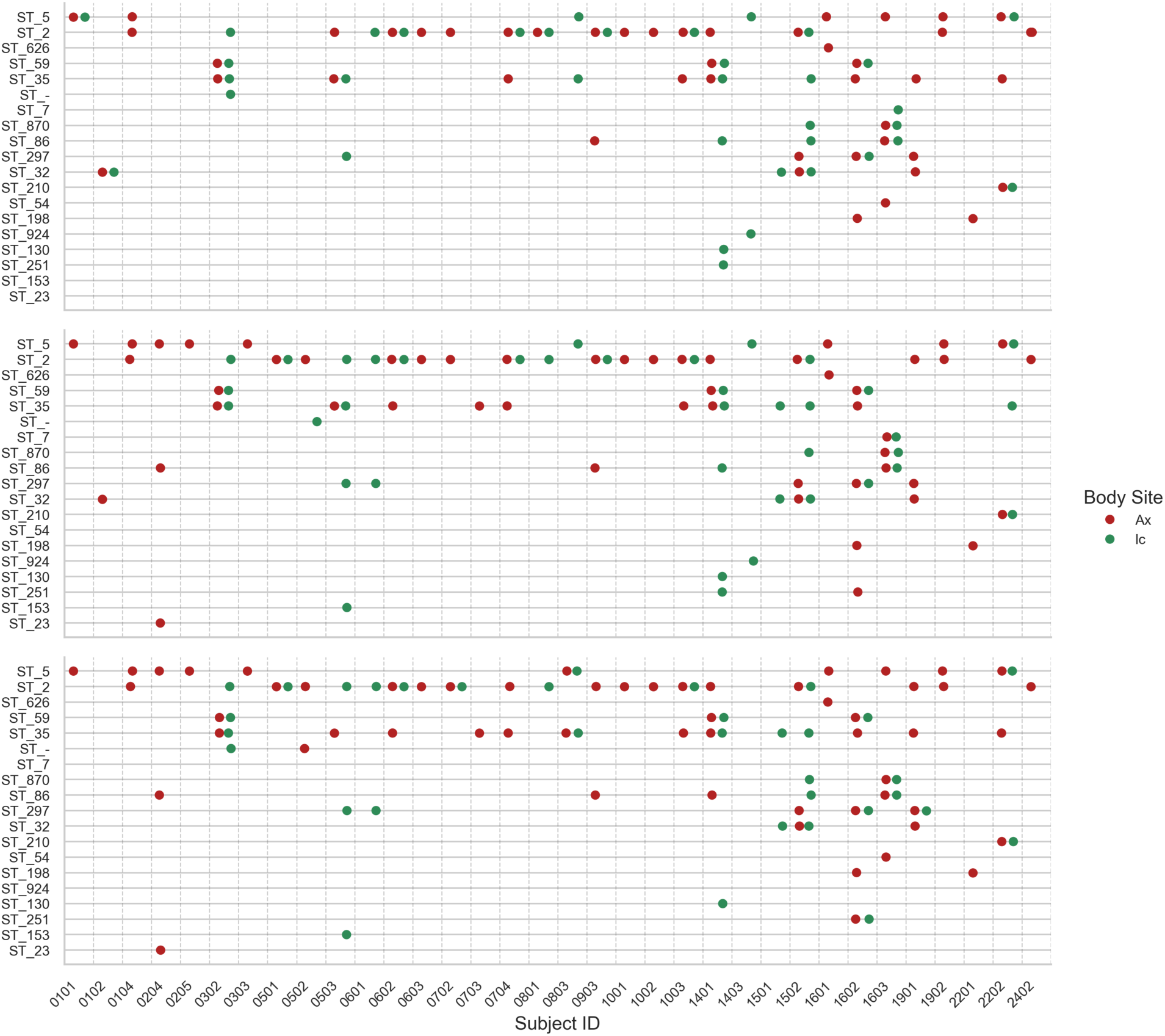
Persistent detection of multiple sequence types of *Staphylococcus epidermidis* by StrainGE in shotgun metagenomic data.

**Figure S6.**
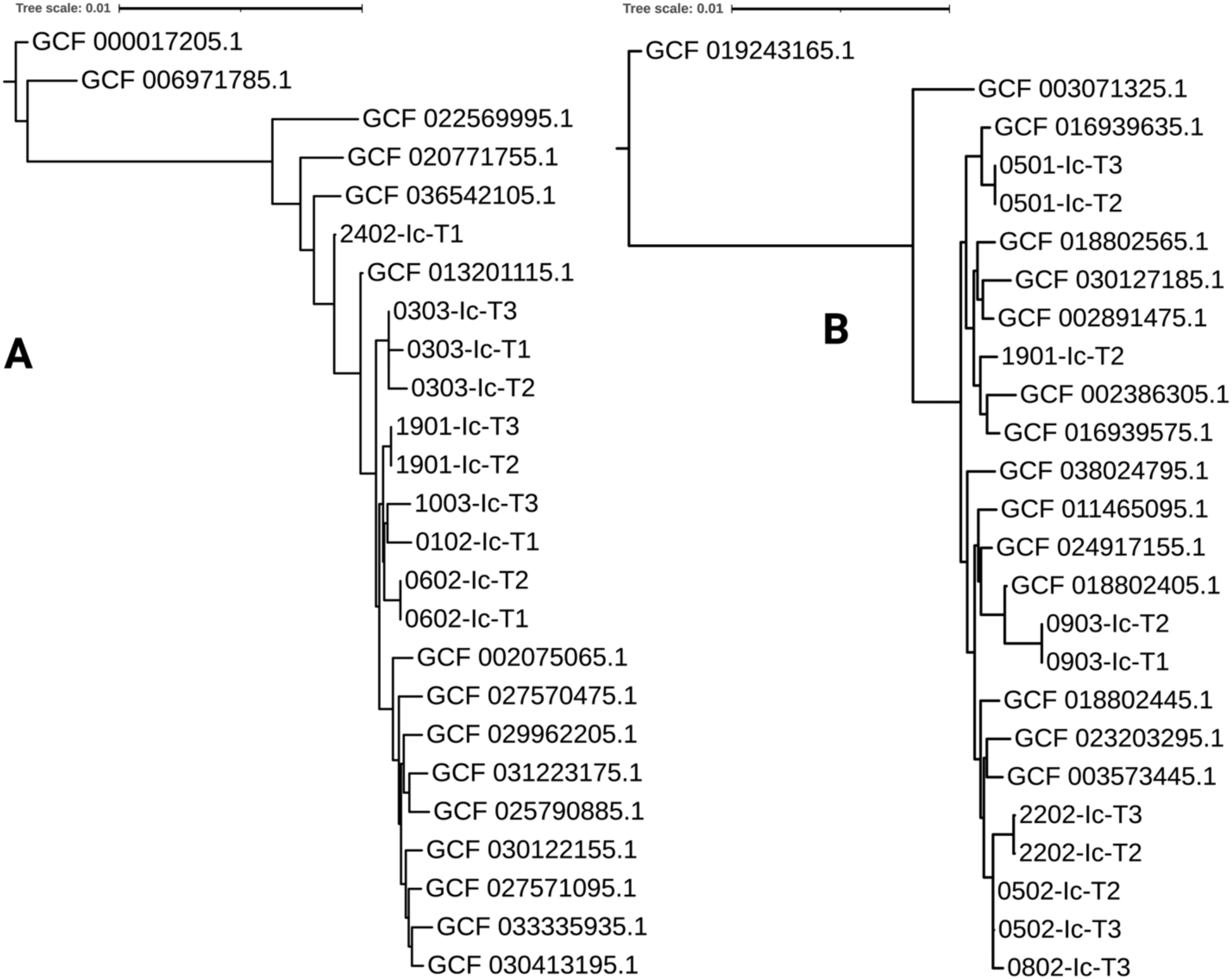
Genomic relatedness of healthcare-associated microbes in nursing home Residents. (A) Phylogenetic tree of *Pseudomonas aeruginosa*. (B) Phylogenetic tree of *Morganella morganii*.

**Figure S7.**
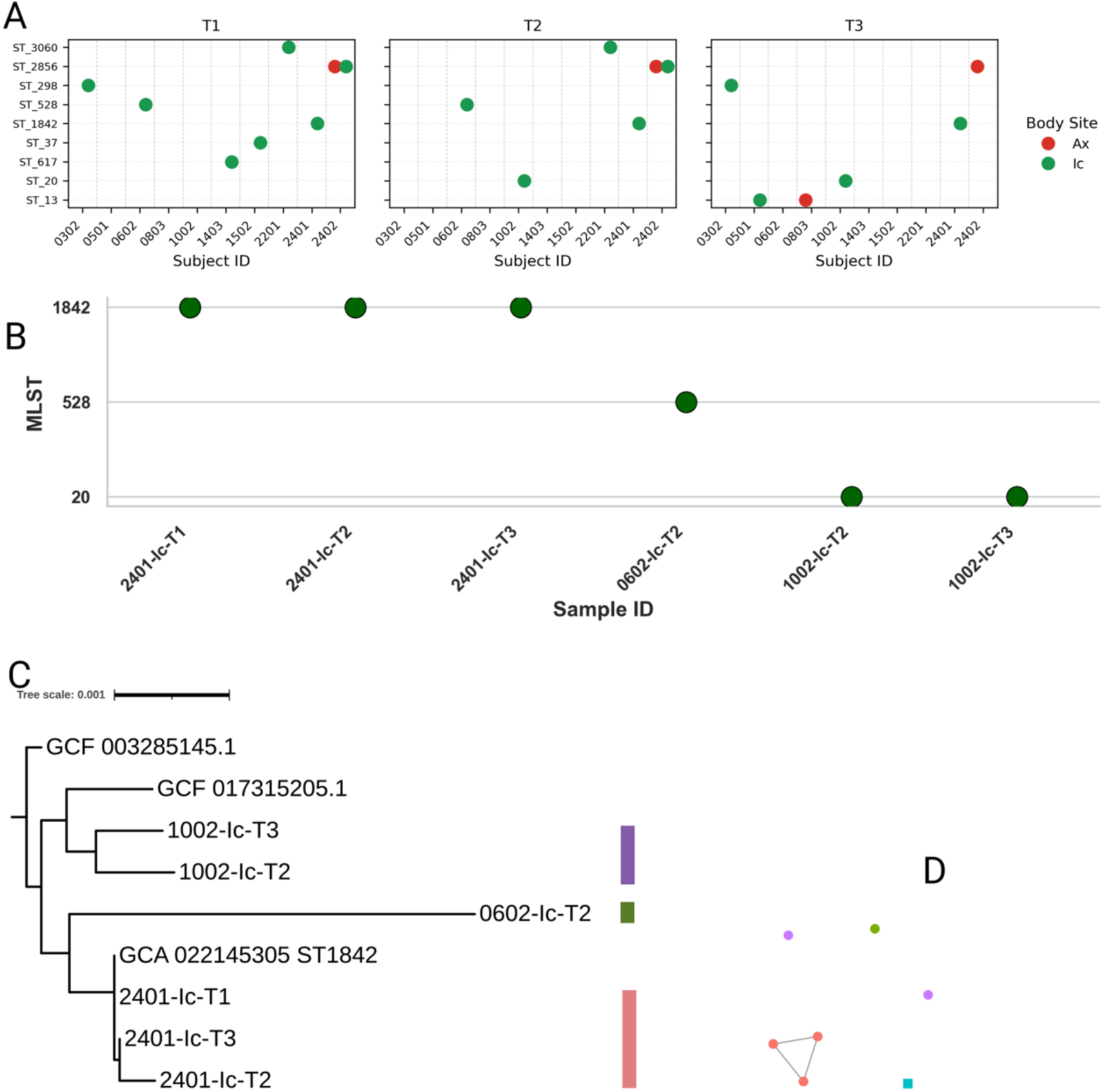
Detection and genomic characterization of *Klebsiella pneumoniae* on NH resident skin. **(A)** Read-level detection of *K. pneumoniae* strains across three timepoints using StrainGE. **(C)** MAG-derived multilocus sequence typing (MLST) for samples with near-complete *K. pneumoniae* genomes. **(C)** Phylogenetic tree of *K. pneumoniae* MAGs from NH residents alongside reference genomes. **(D)** Average nucleotide identity (ANI) network confirming tight clustering of strains within individuals and separation between residents.

**Figure S8.**
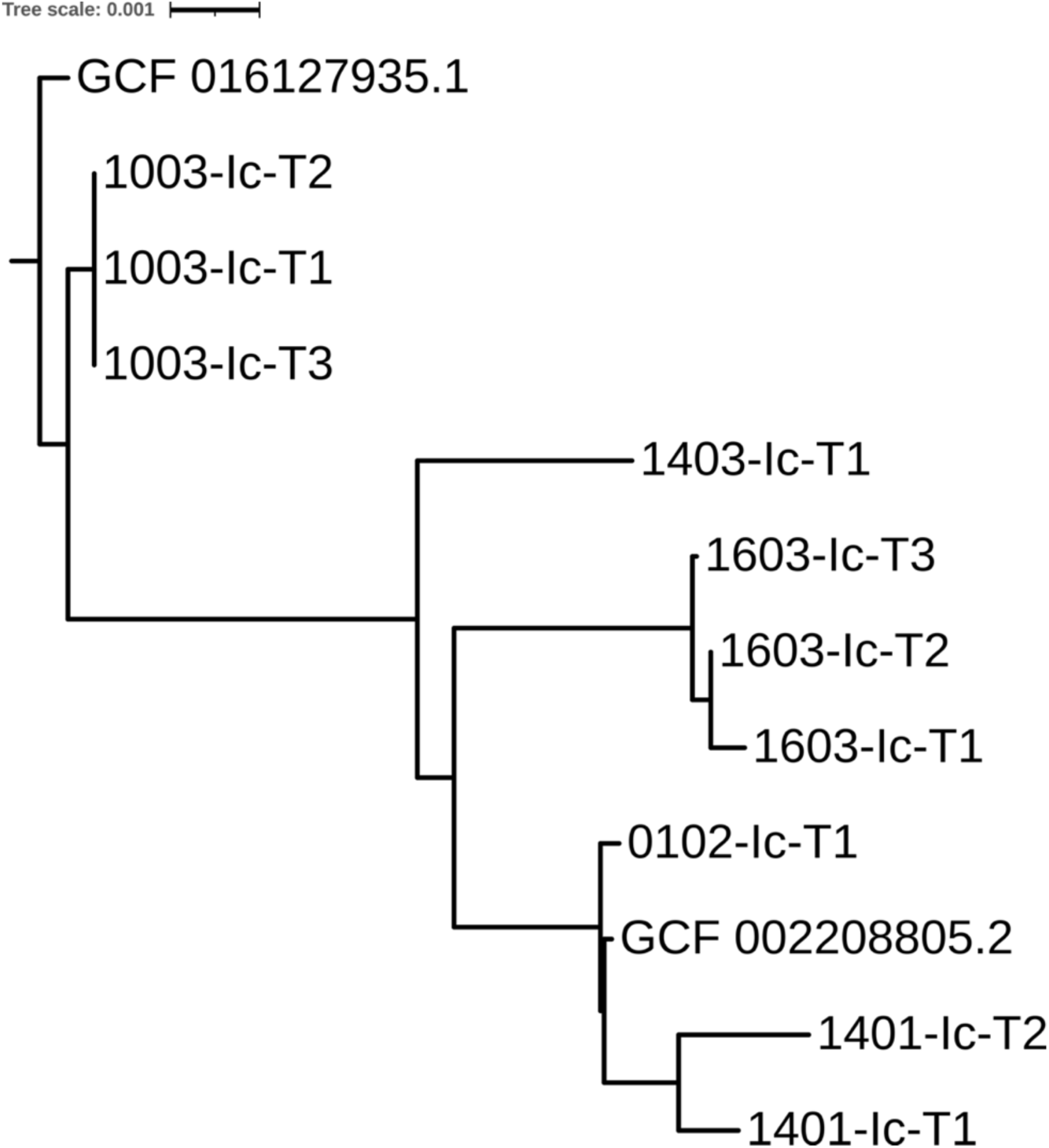
Within-Host Persistence of *Staphylococcus pettenkoferi* Strains in Nursing Home Residents. Single copy marker gene phylogenetic tree of *S. pettenkoferi* MAGs.

